# Metabolic Biomarkers for Peripheral Artery Disease Compared with Coronary Artery Disease: Lipoprotein and metabolite profiling of 31,657 individuals from five prospective cohorts

**DOI:** 10.1101/2020.07.24.20158675

**Authors:** Emmi Tikkanen, Vilma Jägerroos, Rodosthenis Rodosthenous, Michael V Holmes, Naveed Sattar, Mika Ala-Korpela, Pekka Jousilahti, Annamari Lundqvist, Markus Perola, Veikko Salomaa, Peter Würtz

## Abstract

**Background:** Peripheral artery disease (PAD) and coronary artery disease (CAD) represent atherosclerosis in different vascular beds. We conducted detailed metabolic profiling to identify biomarkers for the risk of developing PAD and compared with risk of CAD to explore common and unique risk factors for these different vascular diseases.

**Methods:** We measured blood biomarkers using nuclear magnetic resonance metabolomics in five Finnish prospective general-population cohorts (FINRISK 1997, 2002, 2007, 2012, and Health 2000 studies, n = 31 657). We used Cox modelling to estimate associations between biomarkers and incident symptomatic PAD and CAD (498 and 2073 events, respectively) during median follow-up time of 14 years.

**Results:** The pattern of biomarker associations for incident PAD deviated from that for CAD. Apolipoproteins and cholesterol measures were robustly associated with incident CAD (for example, age- and sex-adjusted hazard ratio per SD for higher apolipoprotein B/A-1 ratio: 1.30; 95% confidence interval 1.25-1.36), but not with incident PAD (1.04; 0.95-1.14; P_heterogeneity_<0.001). Low-density lipoprotein (LDL) particle concentrations were also associated with incident CAD (e.g. small LDL particles: 1.24; 1.19-1.29) but not with PAD (1.07; 0.98-1.17; P_heterogeneity_<0.001). In contrast, more consistent associations of smaller LDL particle size and higher triglyceride levels in LDL and HDL particles with increased risk for both CAD and PAD events were seen (P_heterogeneity_>0.05). Many non-traditional biomarkers, including fatty acids, amino acids, inflammation- and glycolysis-related metabolites were associated with future PAD events. Lower levels of linoleic acid, an omega-6 fatty acid, and higher concentrations of glucose, lactate, pyruvate, glycerol and glycoprotein acetyls were more strongly associated with incident PAD as compared to CAD (Pheterogeneity<0.001). The differences in metabolic biomarker associations for PAD and CAD remained when adjusting for body mass index, smoking, prevalent diabetes, and medications.

**Conclusions:** The metabolic biomarker profile for future PAD risk is largely distinct from that of CAD. This may represent pathophysiological differences and may facilitate risk prediction.

## Introduction

Peripheral artery disease (PAD) is a common atherosclerotic disease affecting body extremities. It can lead to serious complications, including limb ischemia and amputation, and is predictive of future stroke and myocardial infarction. Atherosclerosis is underpinning the pathogenesis in cardiovascular diseases, but studies suggest that PAD risk factors are partly different from cerebrovascular and coronary artery disease (CAD)^1-3^. For example, smoking and diabetes appear to be stronger risk factors, whereas hypertension and dyslipidemia are more modest for PAD compared with CAD^2,3^. Recent genome-wide association studies have identified partly distinct genetic signals for PAD compared to CAD^1^. Thus, uncovering precise metabolic pathways underlying PAD is required to better understand the disease pathology and to improve risk evaluation and treatment.

Large prospective studies have demonstrated the utility of detailed metabolic profiling (also known as metabolomics) in uncovering biomarkers for cardiovascular event risk^4,5^ and elucidating the molecular pathophysiology^6^, which might lead to discovery of new therapeutic targets. The overall pattern of metabolic biomarker associations with a given outcome may also help to dissect similarities and differences between various risk factors^7,8^or diseases^4^. This concept recently demonstrated coherent metabolic signatures for future myocardial infarction and ischemic stroke risk, whereas the metabolic signature for intracerebral hemorrhage risk was partly distinct^4^. Only a few studies have examined biomarker associations with future PAD risk because it requires a large sample size to achieve sufficient endpoints^9^. Recently, Aday et al.^10^ used advanced lipoprotein testing in the Women’s Health Study to identify different association patterns for PAD compared with a composite measure of cardiovascular and cerebrovascular disease, and suggested that primary prevention of PAD should go beyond LDL-C lowering. However, these findings were based on a single cohort of female health professionals and had modest numbers of incident events. Also, as non-lipid pathways seem to play more important roles in PAD pathophysiology as compared to other vascular endpoints, it is essential to study such biomarkers in greater detail to further characterize these associations. Metabolic profiling studies have shown that several amino acids and fatty acids are associated with future CAD and stroke risk more strongly than routine lipids^4,5^, but the associations with PAD risk have yet to be evaluated.

In this study, we aimed to characterize the detailed lipoprotein and metabolite profile for PAD in large prospective cohorts of the general population and compare the results with metabolic biomarker pattern for future CAD events.

## Methods

### Data Availability

The data are available for purposes of reproducing the results and additional research via application to the Finnish Institute for Health and Welfare Biobank (https://thl.fi/en/web/thl-biobank).

### Study populations

This observational study examined lipid and metabolite associations with incident PAD and CAD events in five population-based cohorts conducted by the Finnish Institute for Health and Welfare: the FINRISK 1997, 2002, 2007, and 2012 cohorts, and the Health 2000 Study. The cohort studies were approved by the Coordinating Ethical Committee of the Helsinki and Uusimaa Hospital District, Finland. Written informed consent was obtained from all participants. The field surveys for the FINRISK cohort studies have been conducted every five years since 1972 in Finland^11^. The cohorts included in the present study were initiated in 1997, 2002, 2007, and 2012. Health 2000 was conducted during 2000-2001. Each cohort study is an independent random sample drawn from people aged 25-98 (25–74 in FINRISK, and 30 and over in Health 2000) in the Finnish population. Participation rate to these cohorts have been between 60-70%. The data were collected by self-administered questionnaires and clinical measurements (including e.g. weight, height, blood pressure, blood samples) in each survey. All study participants have been followed using nationwide electronic health registries, including national hospital discharge register, causes-of-death register, and drug reimbursement register. Information on use of lipid-lowering medication and blood pressure treatment was obtained from self reports and the drug reimbursement register. Follow-up has been completed until the end of 2016. Pregnant women were excluded from analyses.

### Disease outcome definition

Disease outcomes were derived from national electronic health registries, which cover all cardiovascular events that have led to hospitalization or death in Finland. The cardiovascular diagnoses in these registers have been validated^12-14^. PAD was defined as the first occurrence of atherosclerosis of native arteries of the extremities, peripheral vascular operations, unspecified PAD and diabetes with circulatory complications (ICD-10 codes: E10.5, E11.5, E12.5, E13.5, E14.5, I70.2, I73.9). CAD was defined as the first occurrence of CAD event during the follow-up, comprising fatal or non-fatal myocardial infarction, cardiac revascularization (coronary artery bypass graft surgery or percutaneous transluminal coronary angioplasty), or unstable angina (ICD-10 codes: I21-I25, I46, R96, R98). To focus on biomarkers for incident disease, individuals with CAD, PAD, or stroke at baseline according to the registry data were excluded from all analyses. Prevalent and incident diabetes status was recorded based on nationwide hospital and medical reimbursement registries.^20^

### Lipid and metabolite biomarker profiling

Venous blood was drawn from non-fasting samples, but with recommended minimum of 4-h fast (median recorded fasting 5h, inter-quartile range 4-6h). The samples were collected and centrifuged at the field survey sites and then transported to the quality-controlled central laboratory, where the serum samples have been stored in -70°C or colder^11^. Lipid and metabolite measures were quantified by high-throughput nuclear magnetic resonance (NMR) metabolomics (Nightingale Health Ltd, Helsinki, Finland; biomarker quantification version 2020). This platform provides simultaneous quantification of routine lipids, lipoprotein subclass profiling with lipid concentrations within 14 subclasses, fatty acid composition, and various low-molecular weight metabolites including amino acids, ketone bodies and gluconeogenesis-related metabolites in molar concentration units. NMR metabolomics was conducted for all serum samples available in the cohorts (comprising ∼90% of participants enrolled) using 350µL aliquots. Biomarkers were quantified independently for each sample without using information from other samples in same well-plate or same cohort. The average success rate of metabolite quantification was 99%. Technological details and epidemiological applications of the Nightingale NMR platform have been reviewed previously^15,16^. The process of this NMR metabolomics technology has received regulatory approval (CE) and 37 biomarkers in the panel have been clinically certified for diagnostic use. **Supplementary Figure 1** illustrates the consistency of biomarkers measured both by clinical chemistry assays (conducted soon after sample collection) and Nightingale NMR (conducted from samples stored 6-15 years prior to NMR measurements), with correlation coefficients in line with earlier reports^4,15,17^. To facilitate visualization, we display results for 57 measures spanning most of the metabolic pathways in the main text; complete results for all the 250 measures quantified by the Nightingale NMR platform are reported in **Supplementary Table**.

### Statistical methods

All metabolic markers were scaled to standard deviation (SD) units to enable comparison of biomarker associations for measures with different units and across wide concentration ranges. Results in absolute concentrations are reported in Supplementary Table. No transformation of the metabolite concentrations was employed.

We used Cox proportional hazard modelling to estimate associations between biomarkers and incident PAD and CAD separately in each cohort. Hazard ratios from each cohort were meta-analyzed using inverse-variance weighted fixed-effect models. Follow-up time was censored at the time of first cardiovascular (i.e. PAD or CAD) event and subsequent events were not included in the analysis. We used two different sets of covariates: our primary analysis was conducted adjusting for sex and age, using age as the time scale (Model 1). In secondary analyses, we additionally adjusted for systolic blood pressure, blood pressure treatment, body mass index, prevalent diabetes, smoking (current vs non-smoker), and lipid-lowering medication (Model 2). The overall concordance of the biomarker association pattern with incident PAD and CAD was summarized using the linear fit of the hazard ratios.^4,7^ We also conducted sensitivity analyses excluding all individuals with prevalent diabetes and those who developed diabetes prior to PAD and CAD events.

To account for multiple testing, we used a threshold of P<0.05/(50*2)=0.0005 for statistical significance, which is based on 50 independent tests in the NMR metabolomics data (number of principal components explaining 99% of variation) and two disease outcomes studied. Statistical analyses were conducted using R version 3.6 (R Foundation for Statistical Computing, Vienna, Austria) and plots made using the ggforestplot package.

## Results

The study included 31 657 participants from five general population cohorts in Finland, all free from cardiovascular disease at baseline. Baseline characteristics of each cohort are shown in **Table 1**. During follow-up (median 14 years; interquartile range 9-15 years; 370 000 person years in total), 498 incident PAD events and 2073 CAD events occurred as the first atherosclerotic event. The median age of the first event was 70 years for both CAD and PAD.

**Table 1.** Baseline characteristics and event numbers in the five cohorts.

|  | FINRISK<br>1997 | FINRISK<br>2002 | FINRISK<br>2007 | FINRISK<br>2012 | Health<br>2000 |
| --- | --- | --- | --- | --- | --- |
| **Number of participants** | 7254 | 7575 | 5550 | 5231 | 6047 |
| **Women, N (%)** | 3678 (51) | 4195 (55) | 2939 (53) | 2765 (53) | 3317 (55) |
| **Age, mean (SD)** | 48 (13) | 48 (13) | 50 (14) | 51 (14) | 54 (15) |
| **Body mass index, mean (SD)** | 26.6 (4.5) | 26.8 (4.7) | 27.1 (4.8) | 27.0 (4.9) | 27.0 (4.6) |
| **Systolic blood pressure, mean (SD)** | 136 (20) | 135 (20) | 136 (20) | 134 (19) | 135 (21) |
| **Total cholesterol, mean (SD)** | 5.5 (1.1) | 5.6 (1.1) | 5.3 (1.0) | 5.3 (1.0) | 6.0 (1.1) |
| **HDL cholesterol, mean (SD)** | 1.4 (0.4) | 1.5 (0.4) | 1.4 (0.4) | 1.5 (0.4) | 1.4 (0.4) |
| **Triglycerides, mean (SD)** | 1.5 (1.0) | 1.4 (0.9) | 1.4 (0.9) | 1.4 (0.9) | 1.6 (1.0) |
| **Lipid-lowering medication, N (%)** | 184 (2.5) | 416 (5.5) | 664 (12) | 683 (13) | 342 (5.7) |
| **Antihypertensive medication, N (%)** | 915 (13) | 1022 (13) | 1064 (19) | 1081 (21) | 962 (16) |
| **Current smoking, N (%)** | 1719 (24) | 1948 (26) | 1159 (21) | 997 (19) | 1264 (21) |
| **Prevalent diabetes, N (%)** | 415 (6) | 393 (5) | 439 (8) | 582 (11) | 274 (5) |
| **Incident CAD, N (%)** | 752 (10) | 435 (6) | 228 (4) | 81 (2) | 577 (10) |
| **Incident PAD, N (%)** | 190 (3) | 99 (1) | 70 (1) | 16 (0) | 123 (2) |
| **Median follow-up time, years** | 18.8 | 13.8 | 8.9 | 3.9 | 15.3 |
| Data are number (%) or mean (standard deviation) when appropriate. HDL, high-density lipoprotein; CAD, coronary artery disease; PAD, peripheral artery disease. |  |  |  |  |  |

### Apolipoproteins and lipids

To best enable comparison of pathophysiological differences in biomarker patterns for PAD and CAD, the primary results are reported with adjustment for age and sex only. Apolipoproteins and cholesterol measures were robustly associated with incident CAD events, but were only weak for incident PAD, with hazard ratios close to unity (**Figure 1**). For example, higher concentrations of apolipoprotein B were associated with increased risk for CAD (HR = 1.23 per 1-SD, 95% confidence interval 1.18-1.28), but not with PAD (1.02; 0.93-1.11; P_heterogeneity_<0.001). A similar pattern of results was observed for very-low-density lipoprotein (VLDL) cholesterol and low-density lipoprotein (LDL) cholesterol measures. Also, higher concentrations of apolipoprotein A-1 was associated with lower risk for CAD (HR = 0.84; 0.80-0.88), but not with PAD (1.00; 0.91-1.10; P_heterogeneity_<0.001). High-density lipoprotein (HDL) cholesterol showed similar results. However, higher concentrations of triglycerides in VLDL, LDL as well as HDL were consistently associated with increased risk for both CAD and PAD (P_heterogeneity_>0.05).

**Figure 1.**
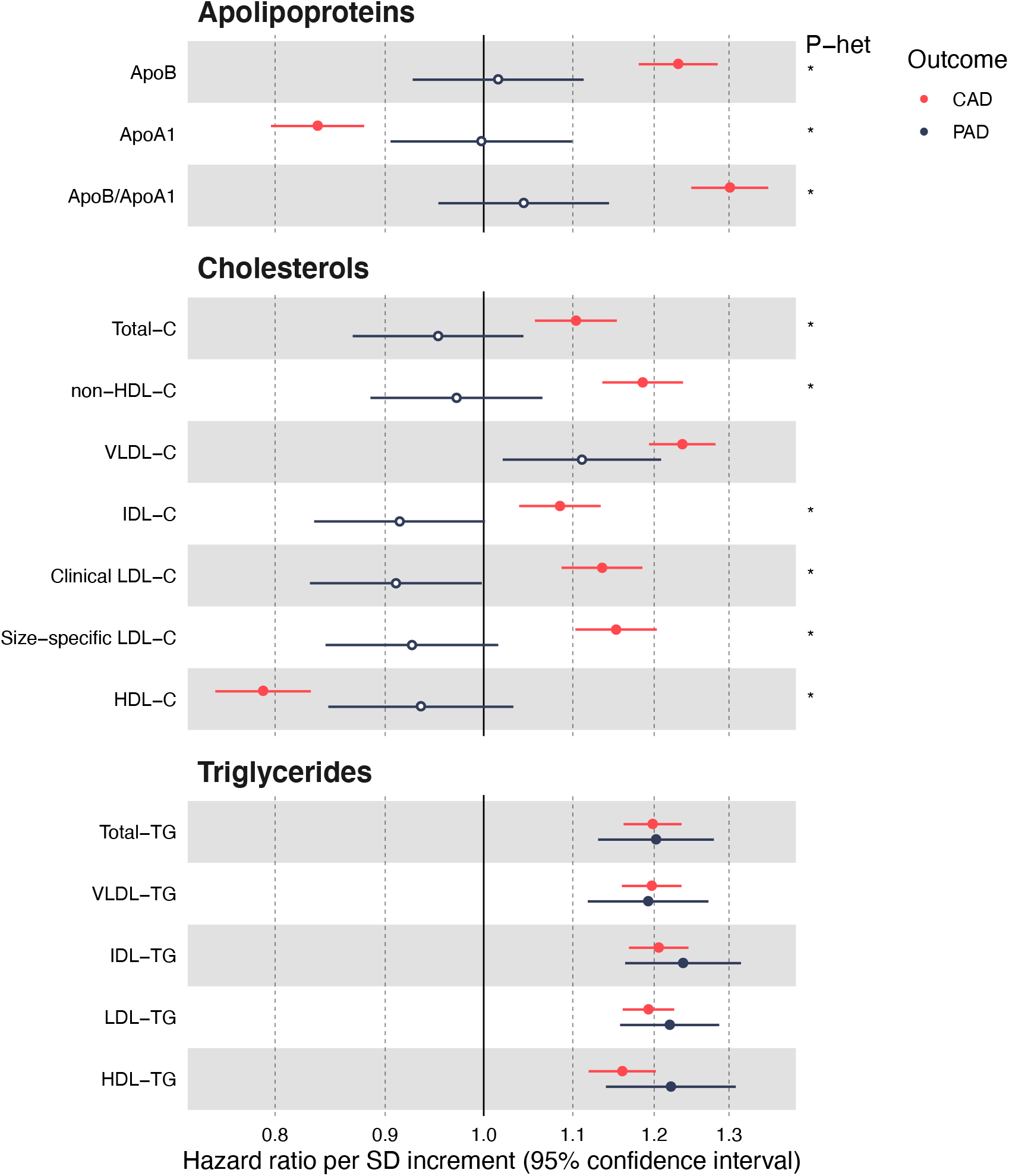
Apolipoprotein and lipid associations with incident CAD (red) and PAD (dark blue). Hazard ratios are per 1-SD higher concentration. Models are adjusted for sex and age. Results are meta-analyzed for 31 657 individuals from five prospective cohorts. Open circles denote P ≥ 0.0005, closed circles P < 0.0005. Asterisks denote P < 0.001 for heterogeneity between PAD and CAD associations. Complete numerical results in absolute concentrations and SD-scaled units are listed in the Supplementary Table. Clinical LDL-C and size-specific LDL-C refer to different methods for defining LDL18. C indicates cholesterol; HDL, high-density lipoprotein; LDL, low-density lipoprotein; TG, triglycerides; VLDL, very low-density lipoprotein.

### Lipoprotein subclasses and particle size

**Figure 2** shows the associations of particle concentrations in 14 lipoprotein subclasses and particle size measures with incident CAD and PAD. For particle concentrations, the associations for LDL and HDL subclasses were generally weak for PAD, except for large VLDL particles. In contrast, almost all lipoprotein particle concentrations were robustly associated with CAD, with the strongest direct associations for small VLDL (1.24, 1.20-1.28) and small LDL (1.24, 1.19-1.29). Also, large and medium-sized HDL particles were inversely associated with incident CAD, whereas they were flat for PAD. Results for specific lipid types within the 14 lipoprotein subclasses broadly followed the same pattern as illustrated for particle concentrations (**Supplementary Figure 2)**. LDL particle size was inversely associated with both endpoints, whereas VLDL particle size and HDL particle size were only associated with incident CAD.

**Figure 2.**
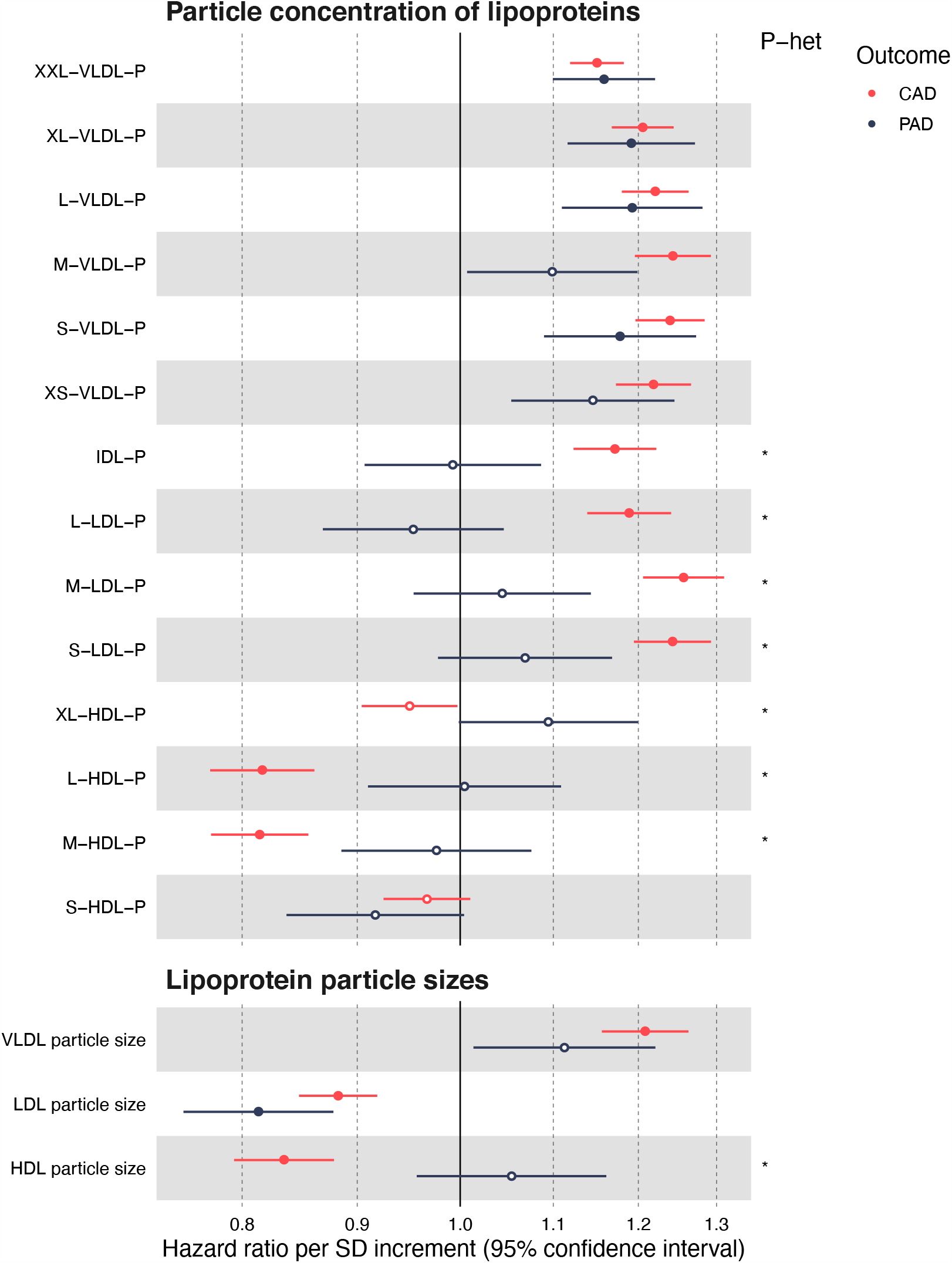
Lipoprotein subclass associations with incident CAD (red) and PAD (dark blue). Hazard ratios are per 1-SD higher concentration. Models are adjusted for sex and age. Open circles denote P ≥ 0.0005, closed circles P < 0.0005. Asterisks denote P < 0.001 for heterogeneity between PAD and CAD associations. HDL indicates high-density lipoprotein; IDL, intermediate-density lipoprotein; LDL, low-density lipoprotein; P indicates particle concentration; VLDL, very low-density lipoprotein.

### Fatty acids, polar metabolites and inflammatory proteins

In addition to lipoprotein and lipid measures, the Nightingale NMR platform simultaneously quantifies a number of fatty acids, polar metabolites and two inflammatory protein measures. Many of these non-traditional biomarkers showed strong associations with both CAD and PAD events (**Figure 3**). The novel biomarkers tended to be particularly strong for PAD, with 11 of the measures having hazard ratios higher than 1.2 or lower than 0.8 per 1-SD. For comparison, the hazard ratio for body mass index is 1.15 (1.05-1.26) per SD in this dataset.

**Figure 3.**
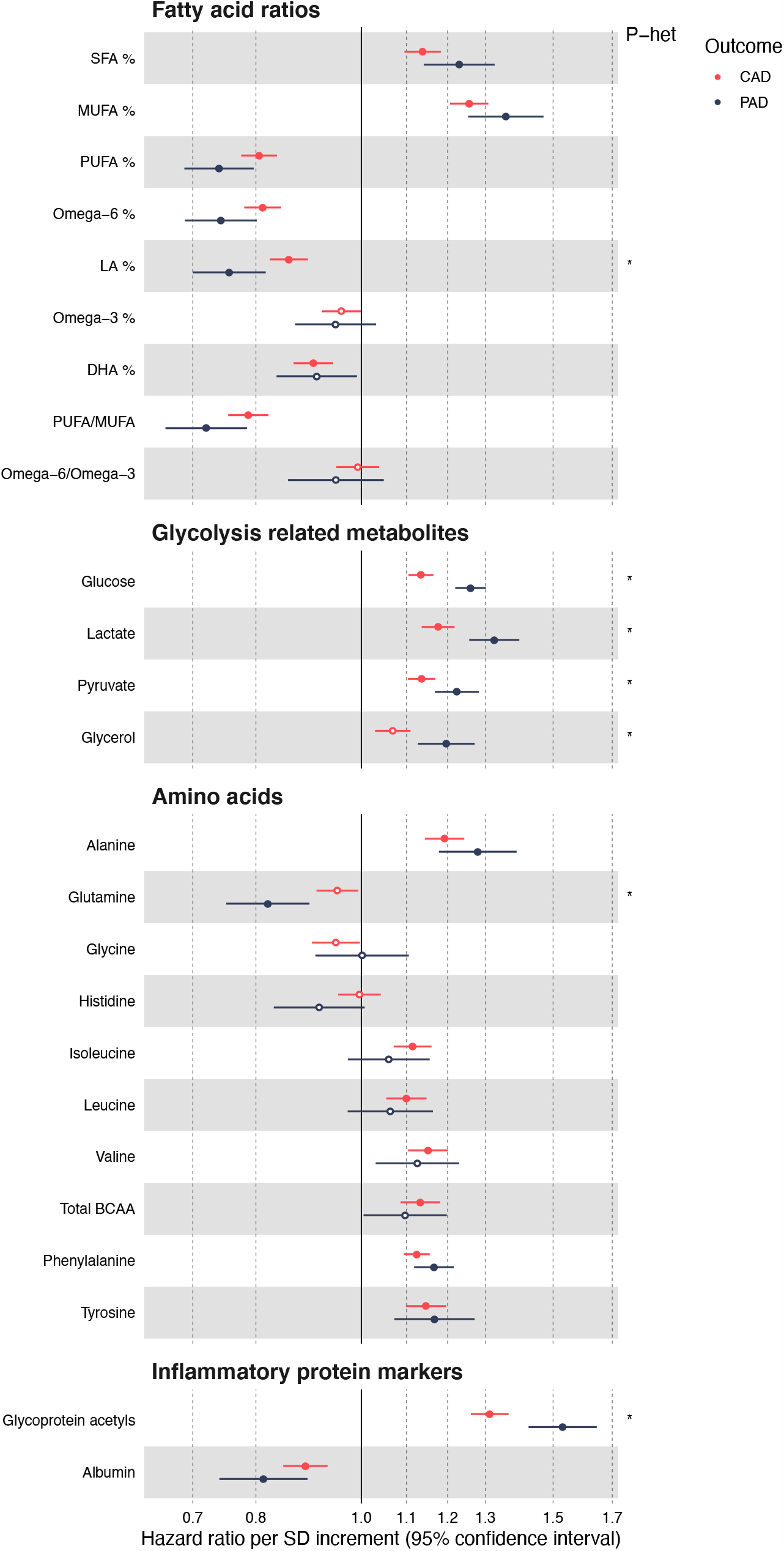
Fatty acid, polar metabolite and inflammatory protein associations with incident CAD (red) and PAD (dark blue). Hazard ratios are per 1-SD higher concentration. Models are adjusted for sex and age. Open circles denote P ≥ 0.0005, closed circles P < 0.0005. Asterisks denote P < 0.001 for heterogeneity between PAD and CAD associations. BCAA indicates branched-chain amino acids; DHA, docosahexaenoic acid; LA, linoleic acid; MUFA, monounsaturated fatty acids; PUFA, polyunsaturated fatty acids; SFA, saturated fatty acids.

For fatty acids, higher proportions of saturated as well as monounsaturated fatty acids (MUFA), relative to total fatty acids, were associated with increased risk for both PAD and CAD. Higher proportions of polyunsaturated fatty acids (PUFA) were associated with decreased risk for both PAD and CAD events, with docosahexaenoic acid (DHA) omega-3 showing similar magnitudes for PAD and CAD, whereas linoleic acid (LA) omega-6 levels had a stronger inverse association with PAD (P_heterogeneity_<0.001).

The four glycolysis-related metabolites measured had stronger associations with PAD than with CAD risk (P_heterogeneity_<0.001), with the strongest associations observed for lactate and pyruvate. For amino acids, the strongest associations for both CAD and PAD were alanine and phenylalanine, whereas glutamine displayed an inverse association with PAD risk. The inflammation-related protein markers albumin and glycoprotein acetyls (GlycA) were associated with both endpoints. GlycA displayed the single strongest hazard ratio per SD-change for both CAD (1.31, 1.26-1.37) and PAD (1.53, 1.42-1.65) risk among all the metabolic biomarkers analysed in this study.

### Biomarker signature for PAD compared with CAD

The overall consistency of the pattern of biomarker associations for PAD and CAD is illustrated in **Figure 4**. The concordance was moderate when comparing the hazard ratios for the 57 highlighted biomarkers (*R*^2^=0.49). The plot reinforces the observation that LDL and HDL lipid biomarkers were only associated with future CAD but not with PAD. For biomarkers predictive of both outcomes, the largest heterogeneity was observed for GlycA and glycolysis-related metabolites, with stronger associations with future PAD events.

**Figure 4.**
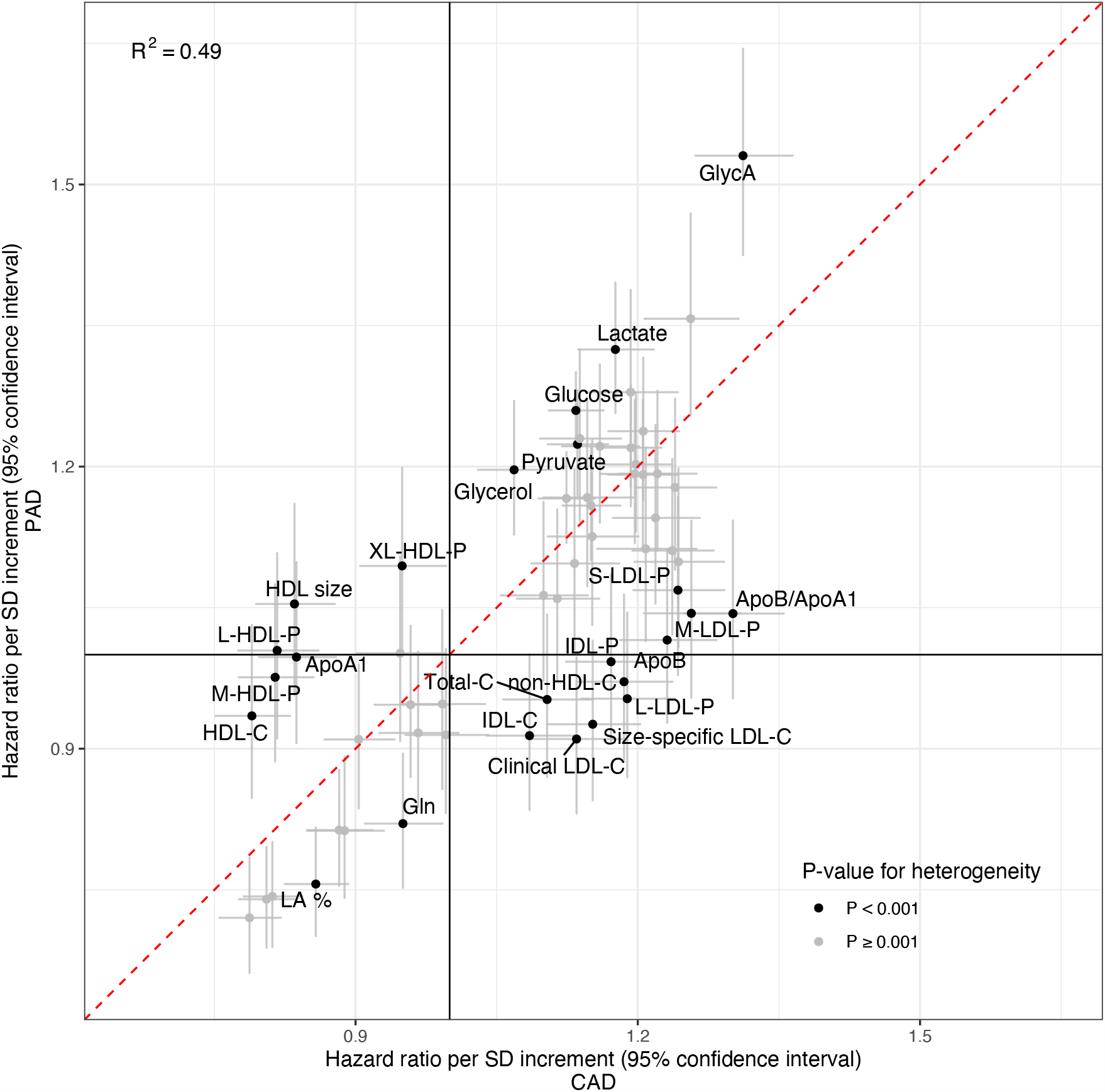
Consistency of metabolic biomarker associations for CAD and PAD. The hazard ratio of each biomarker is given with 95% confidence intervals in gray vertical and horizontal error bars. The red dashed line denotes the diagonal. Biomarkers with P-value < 0.001 for heterogeneity between associations with PAD and CAD are marked by black color coding. PAD indicates peripheral artery disease; CAD, coronary artery disease; GlycA, glycoprotein acetyls; C, cholesterol; HDL, high-density lipoprotein; LDL, low-density lipoprotein; L, large; M, medium; LA, linoleic acid; P, particle concentration; S, small. VLDL, very low-density lipoprotein.

### Complete biomarker associations and sensitivity analyses

Results for all 250 metabolic measures quantified by the Nightingale NMR metabolomics platform are shown in **Supplementary Figure 2**. All numerical results are listed in the **Supplementary Table**. The findings in Figure 1 to 4 are presented as meta-analysis of the five prospective cohorts; the results were highly consistent within individual cohorts for both CAD and PAD, despite large differences in follow-up time and fraction of individuals on lipid-lowering medication at baseline (**Supplementary Figures 3** and **4)**.

We further examined the influence of traditional cardiovascular risk factors on the biomarker associations. The overall biomarker association profiles did not change substantially when further adjusting for systolic blood pressure, body mass index, diabetes status, smoking, and lipid-lowering medication (**Supplementary Figure 5**). In particular, the pronounced differences in CAD and PAD associations for apolipoprotein and cholesterol were similar. Many of the nontraditional biomarkers, such as omega-6 fatty acids, glycolysis metabolites, phenylalanine, albumin, and GlycA, remained significantly associated with future PAD events after adjusting for the traditional risk factors, including prevalent diabetes.

To examine if inclusion of diabetes with circulatory complications in the PAD event definition could partly underpin the observed biomarker differences, we performed sensitivity analysis were all diabetes cases that were registered prior to incidence of PAD and CAD were excluded (**Supplementary Figure 6**). Some of the biomarker associations for PAD were attenuated in this sensitivity analysis (e.g. glucose, alanine, and glutamine), while other biomarker associations with PAD, such as lactate, phenylalanine, and GlycA, remained similar. The differences in apolipoprotein and lipids associations for CAD compared with PAD were unaltered.

Finally, to explore if the biomarker associations with incident PAD was similar those with diabetes, we plotted the biomarker associations with incident diabetes (2490 events). The overall pattern of biomarkers with diabetes was distinct to that for PAD, with stronger associations e.g. for branched-chain amino acids. The lipid biomarker associations with incident diabetes were more similar to those observed for CAD than for PAD (**Supplementary Figure 7**).

## Discussion

Detailed lipoprotein and metabolite profiling in large population-based cohorts uncovered several novel circulating biomarkers reflecting future risk for PAD events. The overall biomarker association profile showed several molecular differences in comparison to biomarkers predictive of future CAD events. We have two main findings: First, the most heterogenous biomarker associations were for standard lipid measures, which were strongly associated with future CAD events but not with the risk for PAD. This difference was particularly marked for apolipoprotein B and LDL cholesterol. Second, a broad range of emerging biomarkers including triglycerides in LDL and HDL particles, circulating fatty acids, glycolysis metabolites, amino acids and inflammatory protein markers showed robust association with both endpoints, but the hazard ratios were generally stronger for future PAD than for CAD. These findings exemplify the possibility for characterization of molecular similarities and differences of related cardiovascular diseases via large-scale metabolic profiling to enhance etiological understanding and potentially help to point towards novel treatment opportunities.

The etiology of atherosclerotic diseases in any arterial bed has historically been considered equivalent, but recently, more detailed lipid and metabolic profiling has revealed partly distinct association profiles for different atherosclerosis subtypes^4,10^. With the aging of the global population and increasing prevalence of obesity and diabetes, it is likely that especially PAD will become more common in future^19^. We identified several novel biomarkers for PAD, which could help to elucidate pathophysiological differences of PAD in relation to other atherosclerotic diseases. For example, our findings suggest that standard lipid testing does not capture the risk for developing PAD. Similar overall conclusions were recently also suggested by Aday et al.^10^ based on standard and advanced lipid testing in the Women’s Health Study. In contrast to Aday et al.^10^, however, we did not observe HDL-C, apolipoprotein A1 or LDL particle concentrations to be associated with future PAD events in our study. These deviating results might be explained by the differences in cohort characteristics and subtle differences in PAD outcome definitions. While the Women’s Health Study only included female healthcare professionals, the participants of the present study is representative of the general Finnish population. Further, our PAD definition was wider compared to theirs, as it also included cases of diabetes mellitus with circulatory complications. However, in our sensitivity analyses that excluded all diabetes cases prior to PAD and CAD events did not alter the prominent differences in lipid associations for CAD compared with PAD. The overall pattern of biomarker associations was also distinct for that observed with incident diabetes.^20^ As an additional novelty, we identified several nonlipid biomarkers to be strongly associated with future PAD events. Many of these novel biomarkers for PAD risk, including inflammation related biomarkers, remained associated in the sensitivity analysis excluding all diabetes cases piror to PAD and CAD events.

There are potential clinical implications of our results. First, our findings of novel risk factors could guide the development of preventative therapies. For example, in light of recent findings from the REDUCE-IT trial on high-dose pure eicosapentaenoic acid (EPA)^21^, omega-3 fatty acids have regained strong interest as treatment target for cardiovascular diseases, and may be suitable for prevention of PAD given benefits on triglyceride pathways. However, our findings are of observational nature, meaning we cannot make deductions on causation. Mendelian randomization analyses may eventually help to provide further information on the etiological roles of omega-3 fatty acids and triglyceride-related pathways in PAD.^1^ Second, while we concluded that standard lipid testing is unlikely to be adequate in PAD risk prediction, we identified novel risk factors for PAD that could potentially improve risk prediction. However, the additive predictive value of these biomarkers needs to be tested in independent datasets.

Our study has strengths and limitations. Strengths include large sample size, spanning five prospective cohorts. This enabled analyses of incident symptomatic PAD and CAD with a high number of events, obtained from nation-wide registries with close to complete coverage of inpatient diagnoses^12-14^. We used a widely-employed high-throughput metabolomics platform that has received regulatory approvals for diagnostics use. The biomarker associations were consistent across all cohorts despite different storage times of samples and non-fasting collection protocols. Furthermore, there was a high consistency between the routine lipids measured by clinical chemistry assays from fresh samples and by NMR 6-15 years after sample collection.

The findings should be interpreted in the context of limitations of this study. First, our PAD diagnosis was based on hospitalized PAD events and thus, does not cover milder forms of PAD. Second, due to observational study settings, causal conclusions cannot be made. However, these results demonstrate the potential of more accurate lipoprotein and metabolic profiling and calls for additional studies to evaluate causality of these biomarkers. Finally, this study was conducted in Finnish population cohorts only. Large prospective studies in other populations are needed to evaluate generalizability and ethnic differences in the biomarker associations.

In conclusion, metabolic profiling of large prospective cohorts highlights prominent differences in the biomarker profile associated with future PAD events compared with CAD events. Whereas routine cholesterol and apolipoprotein measures were not robustly associated with the risk for PAD, metabolic biomarkers reflecting triglyceride metabolism, fatty acid balance, glycolysis, and chronic inflammation were stronger associated with future PAD events than with CAD event. This may suggest that the drivers for atherosclerosis processes are partly distinct in peripheral compared to coronary arteries; however, further studies are needed to determine whether these findings represent meaningful and clinically actionable pathophysiological differences. Our results highlight the need to look beyond atherogenic dyslipidemia for identification of patients at high risk for PAD while still in asymptomatic stages, and for informing development of preventative options and pharmacological treatments for this increasingly prevalent disease.

## Data Availability

The data are available for purposes of reproducing the results and additional research via application to the Finnish Institute for Health and Welfare Biobank.

https://thl.fi/en/web/thl-biobank

https://nightingalehealth.com

## Acknowledgments

The work was primarily funded by Nightingale Health, Ltd. In addition, this research was also supported by Academy of Finland. V. Salomaa was supported by the Finnish Foundation for Cardiovascular Research. M. Holmes works in a unit that receives funding from the UK Medical Research Council and is supported by a British Heart Foundation Intermediate Clinical Research Fellowship (FS/18/23/33512) and the National Institute for Health Research Oxford Biomedical Research Centre. M. Ala-Korpela is supported by a research grant from the Sigrid Juselius Foundation, Finland. We acknowledge study participants for their availability and commitment, and THL biobank for providing the data.

## Disclosures

E. Tikkanen, V. Jägerroos, R. Rodosthenous and P. Würtz are shareholders and/or employees of Nightingale Health, Ltd, a company offering nuclear magnetic resonance based biomarker profiling. V. Salomaa has consulted for Novo Nordisk and Sanofi and received honoraria from these companies. He also has ongoing research collaboration with Bayer AG, all unrelated to this study. M. Holmes has collaborated with Boehringer Ingelheim in research, and in accordance with the policy of The Clinical Trial Service Unit and Epidemiological Studies Unit (University of Oxford), did not accept any personal payment.

## Notes

### Author Declarations

The cohort studies were approved by the Coordinating Ethical Committee of the Helsinki and Uusimaa Hospital District, Finland. Written informed consent was obtained from all participants.

## References

1. Klarin D, Lynch J, Aragam K, Chaffin M, Assimes TL, Huang J, Lee KM, Shao Q, Huffman JE, Natarajan P, Arya S, Small A, Sun YV, Vujkovic M, Freiberg MS, Wang L, Chen J, Saleheen D, Lee JS, Miller DR, Reaven P, Alba PR, Patterson OV, DuVall SL, Boden WE, Beckman JA, Gaziano JM, Concato J, Rader DJ, Cho K, et al. Genome-wide association study of peripheral artery disease in the Million Veteran Program. Nat Med 2019;25:1274–1279.

2. Price JF, Mowbray PI, Lee AJ, Rumley A, Lowe GD, Fowkes FG. Relationship between smoking and cardiovascular risk factors in the development of peripheral arterial disease and coronary artery disease: Edinburgh Artery Study. Eur Heart J 1999;20:344–353.

3. Joshi PH, Martin SS. Unraveling the Risk of Peripheral Artery Disease. Circulation 2018;138:2342–2344.

4. Holmes MV, Millwood IY, Kartsonaki C, Hill MR, Bennett DA, Boxall R, Guo Y, Xu X, Bian Z, Hu R, Walters RG, Chen J, Ala-Korpela M, Parish S, Clarke RJ, Peto R, Collins R, Li L, Chen Z, China Kadoorie Biobank Collaborative Group. Lipids, Lipoproteins, and Metabolites and Risk of Myocardial Infarction and Stroke. J Am Coll Cardiol 2018;71:620–632.

5. Würtz P, Havulinna AS, Soininen P, Tynkkynen T, Prieto-Merino D, Tillin T, Ghorbani A, Artati A, Wang Q, Tiainen M, Kangas AJ, Kettunen J, Kaikkonen J, Mikkilä V, Jula A, Kähönen M, Lehtimäki T, Lawlor DA, Gaunt TR, Hughes AD, Sattar N, Illig T, Adamski J, Wang TJ, Perola M, Ripatti S, Vasan RS, Raitakari OT, Gerszten RE, Casas J-P, et al. Metabolite profiling and cardiovascular event risk: a prospective study of 3 population-based cohorts. Circulation 2015;131:774–785.

6. Cheng S, Shah SH, Corwin EJ, Fiehn O, Fitzgerald RL, Gerszten RE, Illig T, Rhee EP, Srinivas PR, Wang TJ, Jain M, American Heart Association Council on Functional Genomics and Translational Biology; Council on Cardiovascular and Stroke Nursing; Council on Clinical Cardiology; and Stroke Council. Potential Impact and Study Considerations of Metabolomics in Cardiovascular Health and Disease: A Scientific Statement From the American Heart Association. Circ Cardiovasc Genet 2017;10.

7. Würtz P, Wang Q, Kangas AJ, Richmond RC, Skarp J, Tiainen M, Tynkkynen T, Soininen P, Havulinna AS, Kaakinen M, Viikari JS, Savolainen MJ, Kähönen M, Lehtimäki T, Männistö S, Blankenberg S, Zeller T, Laitinen J, Pouta A, Mäntyselkä P, Vanhala M, Elliott P, Pietiläinen KH, Ripatti S, Salomaa V, Raitakari OT, Järvelin M-R, Smith GD, Ala-Korpela M. Metabolic signatures of adiposity in young adults: Mendelian randomization analysis and effects of weight change. Sheehan NA, ed. PLoS Medicine 2014;11:e1001765.

8. Würtz P, Wang Q, Soininen P, Kangas AJ, Fatemifar G, Tynkkynen T, Tiainen M, Perola M, Tillin T, Hughes AD, Mäntyselkä P, Kähönen M, Lehtimäki T, Sattar N, Hingorani AD, Casas J-P, Salomaa V, Kivimäki M, Järvelin M-R, Davey Smith G, Vanhala M, Lawlor DA, Raitakari OT, Chaturvedi N, Kettunen J, Ala-Korpela M. Metabolomic Profiling of Statin Use and Genetic Inhibition of HMG-CoA Reductase. J Am Coll Cardiol 2016;67:1200–1210.

9. Hazarika S, Annex BH. Biomarkers and Genetics in Peripheral Artery Disease. Clin Chem 2017;63:236–244.

10. Aday AW, Lawler PR, Cook NR, Ridker PM, Mora S, Pradhan AD. Lipoprotein Particle Profiles, Standard Lipids, and Peripheral Artery Disease Incidence. Circulation 2018;138:2330–2341.

11. Borodulin K, Tolonen H, Jousilahti P, Jula A, Juolevi A, Koskinen S, Kuulasmaa K, Laatikainen T, Männistö S, Peltonen M, Perola M, Puska P, Salomaa V, Sundvall J, Virtanen SM, Vartiainen E. Cohort Profile: The National FINRISK Study. Int J Epidemiol 2017;47:696–696i.

12. Tolonen H, Salomaa V, Torppa J, Sivenius J, Immonen-Raiha P, Lehtonen A. The validation of the Finnish Hospital Discharge Register and Causes of Death Register data on stroke diagnoses. Eur J Cardiovasc Prev Rehabil 2007;14:380–385.

13. Sund R. Quality of the Finnish Hospital Discharge Register: A systematic review. Scand J Public Health 2012;40:505–515.

14. Pajunen P, Koukkunen H, Ketonen M, Jerkkola T, Immonen-Raiha P, Karja-Koskenkari P, Mahonen M, Niemela M, Kuulasmaa K, Palomaki P, Mustonen J, Lehtonen A, Arstila M, Vuorenmaa T, Lehto S, Miettinen H, Torppa J, Tuomilehto J, Kesaniemi YA, Pyorala K, Salomaa V. The validity of the Finnish Hospital Discharge Register and Causes of Death Register data on coronary heart disease. Eur J Cardiovasc Prev Rehabil 2005;12:132–137.

15. Würtz P, Kangas AJ, Soininen P, Lawlor DA, Davey Smith G, Ala-Korpela M. Quantitative Serum Nuclear Magnetic Resonance Metabolomics in Large-Scale Epidemiology: A Primer on -Omic Technologies. Am J Epidemiol 2017;186:1084–1096.

16. Soininen P, Kangas AJ, Würtz P, Suna T, Ala-Korpela M. Quantitative serum nuclear magnetic resonance metabolomics in cardiovascular epidemiology and genetics. Circ Cardiovasc Genet American Heart Association, Inc; 2015;8:192–206.

17. Tikkanen E, Minicocci I, Hällfors J, Di Costanzo A, D’Erasmo L, Poggiogalle E, Donini LM, Würtz P, Jauhiainen M, Olkkonen VM, Arca M. Metabolomic Signature of Angiopoietin-Like Protein 3 Deficiency in Fasting and Postprandial State. Arterioscler Thromb Vasc Biol 2019;39:665–674.

18. Holmes MV, Ala-Korpela M. What is ‘LDL cholesterol’? Nat Rev Cardiol 2019;16:197–198.

19. Criqui MH, Aboyans V. Epidemiology of peripheral artery disease. Circ Res 2015;116:1509–1526.

20. Ahola-Olli AV, Mustelin L, Kalimeri M, Kettunen J, Jokelainen J, Auvinen J, Puukka K, Havulinna AS, Lehtimäki T, Kähönen M, Juonala M, Keinänen-Kiukaanniemi S, Salomaa V, Perola M, Järvelin M-R, Ala-Korpela M, Raitakari O, Würtz P. Circulating metabolites and the risk of type 2 diabetes: a prospective study of 11,896 young adults from four Finnish cohorts. Diabetologia 2019;62:2298–2309.

21. Bhatt DL, Steg PG, Miller M, Brinton EA, Jacobson TA, Ketchum SB, Doyle RT, Juliano RA, Jiao L, Granowitz C, Tardif J-C, Ballantyne CM, REDUCE-IT Investigators. Cardiovascular Risk Reduction with Icosapent Ethyl for Hypertriglyceridemia. N Engl J Med 2019;380:11–22.

